# MELD Project: Atlas of lesion locations and postsurgical seizure freedom in focal cortical dysplasia

**DOI:** 10.1101/2021.02.01.21250734

**Authors:** Konrad Wagstyl, Kirstie Whitaker, Armin Raznahan, Jakob Seidlitz, Petra E Vértes, Stephen Foldes, Zachary Humphreys, Wenhan Hu, Jiajie Mo, Marcus Likeman, Shirin Davies, Matteo Lenge, Nathan T Cohen, Yingying Tang, Shan Wang, Mathilde Ripart, Aswin Chari, Martin Tisdall, Nuria Bargallo, Estefanía Conde-Blanco, Jose Carlos Pariente, Saül Pascual-Diaz, Ignacio Delgado-Martínez, Carmen Pérez-Enríquez, Ilaria Lagorio, Eugenio Abela, Nandini Mullatti, Jonathan O’Muircheartaigh, Katy Vecchiato, Yawu Liu, Maria Caligiuri, Ben Sinclair, Lucy Vivash, Anna Willard, Jothy Kandasamy, Ailsa McLellan, Drahoslav Sokol, Mira Semmelroch, Ane Kloster, Giske Opheim, Clarissa Yasuda, Kai Zhang, Khalid Hamandi, Carmen Barba, Renzo Guerrini, William Davis Gaillard, Xiaozhen You, Irene Wang, Sofía González-Ortiz, Mariasavina Severino, Pasquale Striano, Domenico Tortora, Reetta Kalviainen, Antonio Gambardella, Angelo Labate, Patricia Desmond, Elaine Lui, Terry O’Brien, Jay Shetty, Graeme Jackson, John S Duncan, Gavin P Winston, Lars Pinborg, Fernando Cendes, J Helen Cross, Torsten Baldeweg, Sophie Adler

**Author notes:** Correspondence to: Dr Konrad Wagstyl, Wellcome Centre for Human Neuroimaging, 12 Queen Square, London WC1N 3AR.

## Abstract

Drug-resistant focal epilepsy is often caused by focal cortical dysplasias (FCDs). The distribution of these lesions across the cerebral cortex and the impact of lesion location on clinical presentation and surgical outcome are largely unknown. We created a neuroimaging cohort of patients with individually mapped FCDs to determine factors associated with lesion location and predictors of postsurgical outcome.

The Multi-centre Epilepsy Lesion Detection (MELD) project collated a retrospective cohort of 580 patients with epilepsy attributed to FCD from 20 epilepsy centres worldwide. MRI-based maps of individual FCDs with accompanying demographic, clinical and surgical information were collected. We mapped the distribution of FCDs, examined for associations between clinical factors and lesion location, and developed a predictive model of postsurgical seizure freedom.

FCDs were non-uniformly distributed, concentrating in the superior frontal sulcus, frontal pole and temporal pole. Epilepsy onset was typically before age 10. Earlier epilepsy onset was associated with lesions in primary sensory areas while later epilepsy onset was associated with lesions in association cortices. Lesions in temporal and occipital lobes tended to be larger than frontal lobe lesions. Seizure freedom rates varied with FCD location, from around 30% in visual, motor and premotor areas to 75% in superior temporal and frontal gyri. The predictive model of postsurgical seizure freedom had a positive predictive value of 70% and negative predictive value of 61%.

FCD location is an important determinant of its size, the age of epilepsy onset and the likelihood of seizure freedom post-surgery. Our atlas of lesion locations can be used to guide the radiological search for subtle lesions in individual patients. Our atlas of regional seizure freedom rates and associated predictive model can be used to estimate individual likelihoods of postsurgical seizure freedom. Data-driven atlases and predictive models are essential for evidence-based, precision medicine and risk counselling in neurology.

## Introduction

Epilepsy is one of the most common neurological conditions, with a lifetime risk of 1 in 26.^1^ Focal cortical dysplasia (FCD) is a malformation of cortical development and a common cause of drug-resistant epilepsy.^2,3^ In many patients, FCD is amenable to surgical resection, with reported long term seizure freedom rates of 69%.^4^

FCDs can be categorised into distinct histopathological subtypes.^2^ FCD Type I is characterised by cortical dyslamination, FCD Type II by dysmorphic neurons and dyslamination, and is subdivided into IIa and IIb, with the latter having balloon cells. FCD Type III is associated with another principal lesion, e.g. hippocampal sclerosis.

FCDs can occur anywhere in the cerebral cortex, but different histopathological subtypes show some lobar specificity.^2,3^ FCD Type II lesions are more frequently found in the frontal lobe, whereas FCD Type I and III are more frequently located in the temporal lobe. To date, most studies analysing localisation have used coarse lobar categorisations^4^ or have been limited by small sample sizes^5^. Despite anatomical mapping of lesions being available using presurgical MRI^6–8^ and the emergence of large collaborative neuroimaging studies,^9^ these techniques have not been used to map the topography of FCDs.

The gold-standard treatment for drug-resistant focal epilepsy is surgical resection.^10^ However, a significant proportion of patients (31% of FCD Type II and 42% of FCD Type I)^4^ continue to have seizures post-operatively. Identifying factors relating to seizure freedom is important. Some can be modified to improve patients’ clinical and surgical treatment. Others can be incorporated into machine-learning models to produce patient-specific predictions of seizure freedom for use in clinical planning and risk counselling. Across all focal epilepsies, duration of epilepsy, age at surgery, lesion lobe and histopathological diagnosis are predictors of postsurgical freedom.^4^ Within FCD, the most consistent predictive factors include complete resection of the FCD,^11,12^ temporal resections,^13,14^ having an MRI-visible lesion^15^ and the underlying histopathology being FCD Type II.^4^ However, the relationship between precise lesion location and seizure freedom is unknown.

Detailed spatial mapping of FCD lesions would broaden our understanding of this disease, enabling linkage of a patient’s lesion location to presurgical clinical information. To this end, we created the Multi-centre Epilepsy Lesion Detection (MELD) project to collate a large neuroimaging cohort of patients, including MRI lesion maps with demographic, clinical and surgical variables. We aimed to 1) map the topographic distribution of epileptogenic FCDs across the cerebral cortex, 2) identify clinical factors associated with lesion location and 3) establish predictors of postsurgical seizure freedom.

## Materials and methods

### 1. MELD project consortium

We established the MELD project (https://meldproject.github.io/) involving 20 research centres across five continents. Each centre received approval from their local institutional review board or ethics committee. Written informed consent was provided according to local requirements.

### 2. Participants

Patients over age three were included if they had a 3D T1-weighted MRI brain scan (1.5T or 3T) and a radiological diagnosis of FCD or were MRI-negative with histopathological confirmation of FCD. Participants were excluded if they had previous surgeries, large structural abnormalities in addition to the FCD or T1 scans with gadolinium-enhancement. Centres and patients were given pseudo-anonymised ID codes.

### 3. Site-level data collection and post-processing

#### MRI data collection and post-processing

T1-weighted MRI scans were collected at the 20 participating centres for all participants and cortical surfaces were reconstructed using *FreeSurfer* (versions 5.3 or 6).^*16*^ All MRI data was preoperative in operated patients. Detailed protocols for structural MRI post-processing were adapted from openly available ENIGMA-epilepsy protocols.^9^

#### FCD lesion masking

FCD lesions were delineated on the T1-weighted MRI scans at each site according to our lesion masking protocol.^17^ Lesions were masked by a neuroradiologist or experienced epilepsy researcher at each site. For patients with a radiological diagnosis of FCD, a volumetric lesion mask was created using the T1 scan and 3D FLAIR (where available). For MRI-negative patients but with histopathological confirmation of FCD, the post-operative scan and other corroborative evidence, such as EEG and PET, were used to create the volumetric lesion mask. Volumetric lesion masks were mapped to cortical reconstructions and small defects were filled in using five iterations of a dilation-erosion algorithm. Patients’ lesions were registered to a bilaterally symmetrical template surface, *fsaverage_sym*. Lesion size was calculated as the percentage of lesional vertices out of the total number of vertices in that hemisphere.

#### Participant demographics

The following data were collected for all patients: age at MRI scan, sex, age of epilepsy onset, duration of epilepsy (time from age of epilepsy onset to age at MRI scan), ever reported MRI-negative and histopathological diagnosis (ILAE three-tiered classification system),^2^ seizure-freedom (Engel class I, minimum follow-up time 1 year) and follow-up time in operated patients.

Participant demographic information and lesion masks were shared with the study coordinators for multi-centre analysis. At the point of retrieval by the study coordinators, all participant data were deidentified, as keys linking pseudo-anonymised IDs to patient-identifiable information were stored securely by individual sites.

### 4. Location of focal cortical dysplasias in the cerebral cortex

Lesion masks from all patients were overlaid on the left hemisphere of the template cortical surface to visualise the distribution of lesions for all FCDs, as well as for histopathological subtypes (see Methods Section 5 for tests of interhemispheric asymmetry). Additional surface-based lesion maps for left and right hemispheres were generated for the whole cohort, patients who had and those who had not undergone resective surgery.

Lobar categorisations of lesion location were created based on the lobe which contained the most lesional vertices. For statistical comparisons of lobar location, lesions primarily located in the smallest lobes in the parcellation (cingulate and insula) were assigned to a second lobe (e.g. frontal) which the lesion mask also overlapped. A mask of eloquent cortex was created including the following cortical areas bilaterally from the desikan killiany atlas: precentral (motor), pericalcarine, lateral occipital, cuneus and lingual labels (all visual cortical areas). Additionally the pars opercularis, pars triangularis and transverse temporal labels (language areas) were included on the left hemisphere.

For the creation of a 3D lesion likelihood atlas, aggregated surface-based lesion map values were normalised to between 0 and 1, where the location with the highest number of FCDs had a value of 1. This lesion likelihood atlas was mapped back to the template MRI volume for use as a potential clinical aid in guiding radiological diagnosis.

Bootstrap reproducibility was used to assess the consistency of the overall spatial distribution of FCDs. FCD frequency maps from subsets of 20 to 250 patients were correlated with the map from a subset of 250 patients. This process was randomised and repeated 1000 times. Unstable maps had low correlations and indicated that the sample size was too small. A predictive learning curve was fitted to the R values of different subcohort sizes and interpolated to determine the sample size required for a consistent spatial distribution in the lesion pattern.^18^

### 5. Factors associated with lesion distribution

We trained logistic regression models to test for associations between lesion location and clinical data (Supplementary Fig. 1). The models were fitted to assess which preoperative factors are associated with the presence or absence of lesions at a particular vertex (a point on the cortical surface). The following preoperative factors were included: sex, age of epilepsy onset, duration of epilepsy, ever reported MRI-negative, lesion size and lesion hemisphere. Due to collinearity with duration of epilepsy (*r* = 0.76), age at MRI scan was tested in a separate model excluding duration. Regression coefficients were tested for significance against those calculated on 1000 randomly permuted cohorts. Factors were deemed significant if their coefficient was less than 2.5% or greater than 97.5% of the coefficients from permuted cohorts. Approximately 5% of vertices would be significant by chance. A given factor was considered significantly related to lesion location if the number of significant vertices exceeded the 100-(5/*n*th) percentile number of significant vertices from the permuted cohorts, where n was the number of factors being tested.

Post-hoc analysis of significant factors included testing for similarity between the surface-based map of age of epilepsy onset and a potential explanatory variable, the principal gradient of functional organisation from primary to association areas^19^. To account for spatial autocorrelation, statistical significance was established by comparing correlations with those from 1000 spherically rotated maps.^20^

### 6. Factors associated with seizure freedom

Using the cohort of operated FCD patients with seizure outcome, we calculated the proportion of patients who were seizure free with lesions at each vertex. To assess the association of lesion location with postsurgical seizure freedom, along with other clinical factors, three logistic regression models were fitted. The first was solely based on lesion location, predicting seizure freedom at each cortical vertex based on which patients had lesions overlapping that vertex. A second logistic regression model was fitted to predict seizure freedom using the following presurgically available factors: duration of epilepsy, age of epilepsy onset, MRI-negative status, scanner, lesion overlap with eloquent cortex and lesion size. Due to collinearity with duration of epilepsy, age at MRI scan was tested in a separate model excluding duration. A third model was created including the post-operatively determined histopathological FCD subtype and an interaction between lesion size and histopathological subtype. We calculated the predicted percentage likelihood of seizure freedom from each logistic regression model for each patient. Statistical significance for these regression models was established using the same permutation procedures as in Methods Section 5.

Finally, to assess the predictive value of the presurgical factors (model 2) in determining postsurgical seizure freedom, we carried out 10-fold cross-validation of the model. Sensitivity, specificity, positive predictive value and negative predictive value were calculated.

### 7. Comprehensive search for inter-relationships between demographic, lesional and surgical variables

Relationships between demographic, lesion and surgical variables were systematically analysed (Fig. 4B). Heavily skewed variables were normalised for statistical testing using a Box-Cox transform. The Benjamini-Hochberg procedure was used to control the false discovery rate when conducting multiple comparisons, with *α* set to 0.05.^21^

### 8. Code

All data analysis was performed in Python using the following packages: NumPy, scikit-learn, scipy, pandas, nibabel, matplotlib, seaborn, PtitPrince. All protocols and code for MELD preprocessing and Prediction Of Outcome & Location (POOL) is available to download from <u>protocols.io/researchers/meld-project</u> and www.github.com/MELDProject.

## Data availability

Lesion maps for the whole cohort are freely available to download from the UCL data repository. Code to reproduce all analyses and figures in this manuscript is available to download from github.com/MELDproject. Requests can be made for access to the full MELD dataset.

## Results

### 1. Participant demographics

Data was collected from 580 FCD patients. Demographic information is available in Table 1. Histopathological diagnosis was available in 380 patients (66%, Table 2). Postsurgical outcome data with 1 year minimum follow up was available in 275 patients (47% of operated patients); 65% were seizure free (Engel class 1) at last follow-up (median follow up = 2 years). Seizure freedom rates across histopathological subtypes are presented in Table 2. Although FCD Type I had a lower mean seizure freedom (55%) than FCD Type IIA (69%) and FCD Type IIB (66%), there was no significant difference in outcome according to histopathological subtype.

**Table 1:** Demographics Table

|  | FCD patients (n=580) |
| --- | --- |
| Age at scan (median, IQR) | 19·0 (11·0 - 31·3) |
| Sex (female:male) | 281:298 |
| Age at onset (years) | 6·0 (2·5 - 12·0) |
| Ever reported MRI negative | 188/580 (32%) |
| Duration of epilepsy (years) | 10·4 (4·9 - 19·0) |
| Surgery performed | 423/580 (73%) |
| Histopathology available | 380/580 (66%) |
| Outcome available | 275/580 (47%) |
| Follow up time (median, IQR) | 2·0 (1·3 - 3·4) |

**Table 2:** Histopathology and surgical outcome

| FCD subtype | Histopathology (n, %) | Seizure free (n, %) |
| --- | --- | --- |
| All | 380 (100%) | 165/252 (65%) |
| FCD Type I | 42 (11%) | 17/31 (55%) |
| FCD Type IIA | 118 (31%) | 58/84 (69%) |
| FCD Type IIB | 199 (52%) | 80/121 (66%) |
| FCD Type III | 21 (6%) | 10/16 (62%) |

*Histopathology = histopathological diagnosis; Histopathology % = percentage of patients with particular histopathological diagnosis out of the total number of patients with histopathology; Seizure free = number of patients with a particular histopathological diagnosis who were seizure free out of the total number of patients with FCD subtype and outcome data available; n= number of patients.*

### 2. Location of focal cortical dysplasias in the cerebral cortex

548 patients had lesion masks (T1w MRI 1.5T *n* = 98, 3T *n* = 450, FreeSurfer v5.3 *n* = 401, v6 *n* = 147). The 32 patients that did not have lesion masks were excluded from subsequent analyses. FCDs were evenly distributed between left and right hemispheres (L:R 266:282). Lesions were located throughout the cortex (262 frontal, 158 temporal, 90 parietal, 20 occipital, 10 cingulate, 8 insula). The MRI scans of 188 patients (32%) had at some point been reported as MRI negative, with a similar proportion of patients scanned on 1.5T or 3T MRI being reported as MRI negative. For visualisation, all lesions were mapped to the left hemisphere of *fsaverage_sym* (Fig. 1A). The distribution of lesions was non-uniform, with propensity for FCDs in the superior frontal sulcus, frontal pole, temporal pole and superior temporal gyrus (Fig. 1A & B). Lesion maps were unstable at sample sizes typical for neuroimaging cohorts of FCD, but improved as sample size increased (Fig. 1C). A predictive learning curve determined that a sample size of around *n* = 400 would be required for a stable distribution of lesions across the cortex. This provided support that our cohort (*n* = 548) is sufficient to be representative.

**Figure 1.**
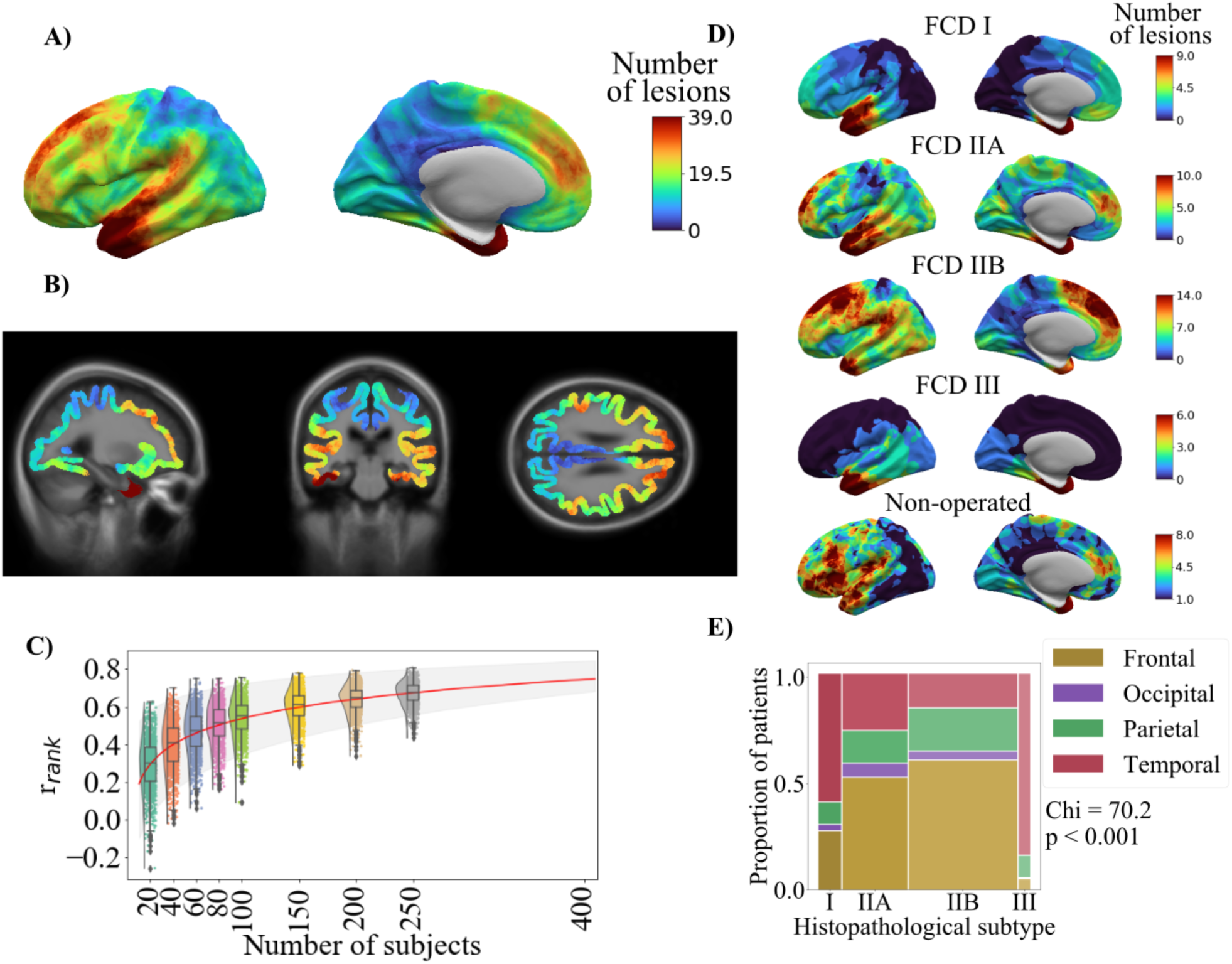
Distribution of FCD lesions across the cerebral cortex. (**A**) All FCD lesion masks mapped to the left hemisphere of the template cortical surface. The distribution of FCDs across the cerebral cortex is non-uniform and with higher concentrations in the superior frontal sulcus, frontal pole, temporal pole and superior temporal gyrus. (**B**) 3D lesion likelihood atlas. Aggregated surface-based lesion map values were normalised to between 0 and 1 and mapped back to the template MRI volume. (**C**) Sample size required for consistent FCD lesion map. Rank correlation (*y* axis) was calculated by comparing the lesion map from a smaller cohort to a larger withheld cohort (*n* = 250). *r*_*rank*_ increased with sample size. Predictive learning curve (red line) estimated that a stable map of lesion distribution requires a sample size of *n* = 400. (**D**) Distribution of FCD lesions according to histopathological subtype. (**E**) Distributions of lesions across cortical lobes within each FCD histopathological subtype. The width of bars indicates the relative numbers of patients. Temporal lobe lesions made up larger proportions of FCD Types I and III, while frontal lobe lesions were more likely to be FCD Types IIA and IIB.

Lesion maps of histopathological subtypes (Fig. 1D) showed that the distribution of FCD lesions across the cortex differs according to histopathological subtype, with a greater proportion of FCD Types I and III lesions located in the temporal lobe (Fig. 1E). In the group of non-operated patients, lesions were most frequently located in the left inferior frontal gyrus and bilateral insula (Fig. 1D, Supplementary Fig. 2). While numbers in this cohort were relatively small, this may represent deliberate avoidance of surgery in language areas and/or challenges in the diagnosis or surgical planning of insula lesions.

### 3. Relationships between demographic variables and lesion distribution

Age of epilepsy onset, duration of epilepsy and lesion size were significantly related to lesion location (number of significant vertices > expected by chance *p <* 0.01) (Fig. 2). Hemisphere, sex and ever reported MRI-negative were not. The age of onset map (Fig. 2A) was significantly correlated with the principal gradient of functional cortical organisation. Lesions in primary areas were associated with a younger age of onset, while association areas had older ages of onset (*r*_*rank*_ = 0.39, *p*_*spin*_ < 0.01). Overall, lesions in temporal, parietal and occipital cortices were associated with a shorter duration of epilepsy (Fig. 2B), whereas lesions around the central sulcus and frontal lobe were associated with a longer duration of epilepsy. This pattern closely resembled the distribution of lesion size and age at scan (Fig. 2C&D), whereby cortical areas associated with longer durations like the frontal lobe tended to have smaller lesions (*r*_*rank*_ = −0.42, *p*_*spin*_ < 0.05) and patients tended to have been older at MRI scan (*r*_*rank*_ = 0.92, *p*_*spin*_ < 0.001).

**Figure 2.**
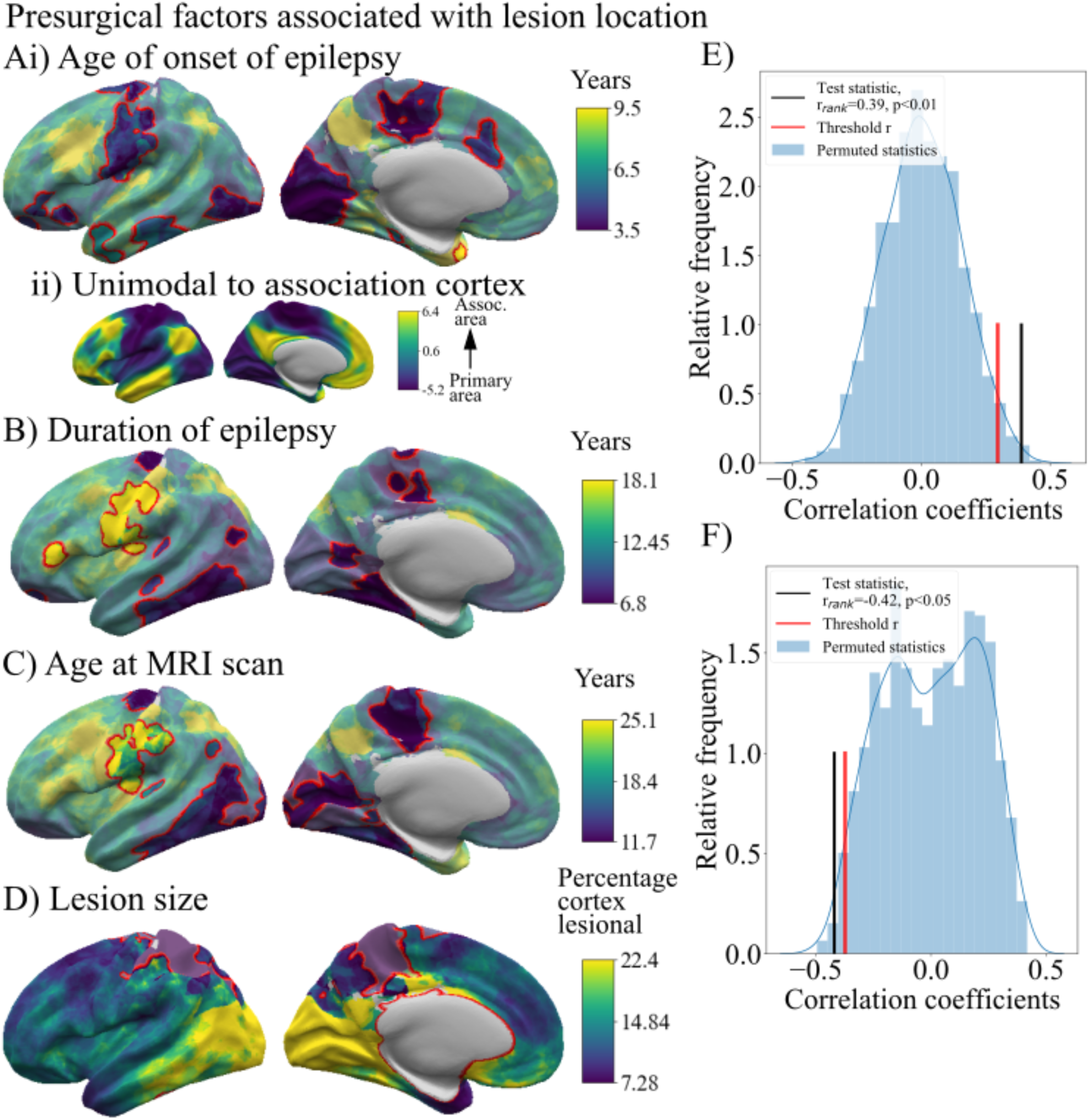
Presurgical factors associated with lesion location. Surface-based maps showing distribution of demographic variables according to lesion location. Vertices outlined in red are significant after comparison with 1000 randomly permuted cohorts. Factors significantly (*p* < 0.01) with lesion location were (**Ai**) age of epilepsy onset, (**B**) duration of epilepsy, (**C**) age at MRI scan and (**D**) lesion size. (**E**) Correlation between the primary gradient of functional organisation (**Aii**) and the age of epilepsy onset (**Ai**) maps in comparison to spatially permuted maps. Lesions in primary areas were associated with a younger age of onset, while association areas had older ages of onset (*r*_*rank*_ = 0.39, *p*_*spin*_ < 0.01). (**F**) Correlation between the duration of epilepsy (**B**) and lesion size (**D**) maps in comparison to spatially permuted maps. Mean duration was significantly negatively correlated with the size of epilepsy lesion, where cortical areas with smaller lesions, e.g. precentral and frontal areas, were associated with a longer duration of epilepsy, whereas areas with larger lesions, e.g. occipital cortex, had shorter durations of epilepsy (*r*_*rank*_ = −0.42, *p*_*spin*_ < 0.05).

### 4. Factors associated with seizure freedom

The percentage of patients who were seizure free post-surgery varied according to lesion location and, therefore, the area of cortex resected (Fig. 3A). The first logistic regression model, based solely on vertex-wise lesion location, showed that visual, motor and premotor areas were associated with significantly lower seizure freedom rates (30-40% of patients were seizure free), likely reflecting conservative resection around eloquent cortex.

**Figure 3.**
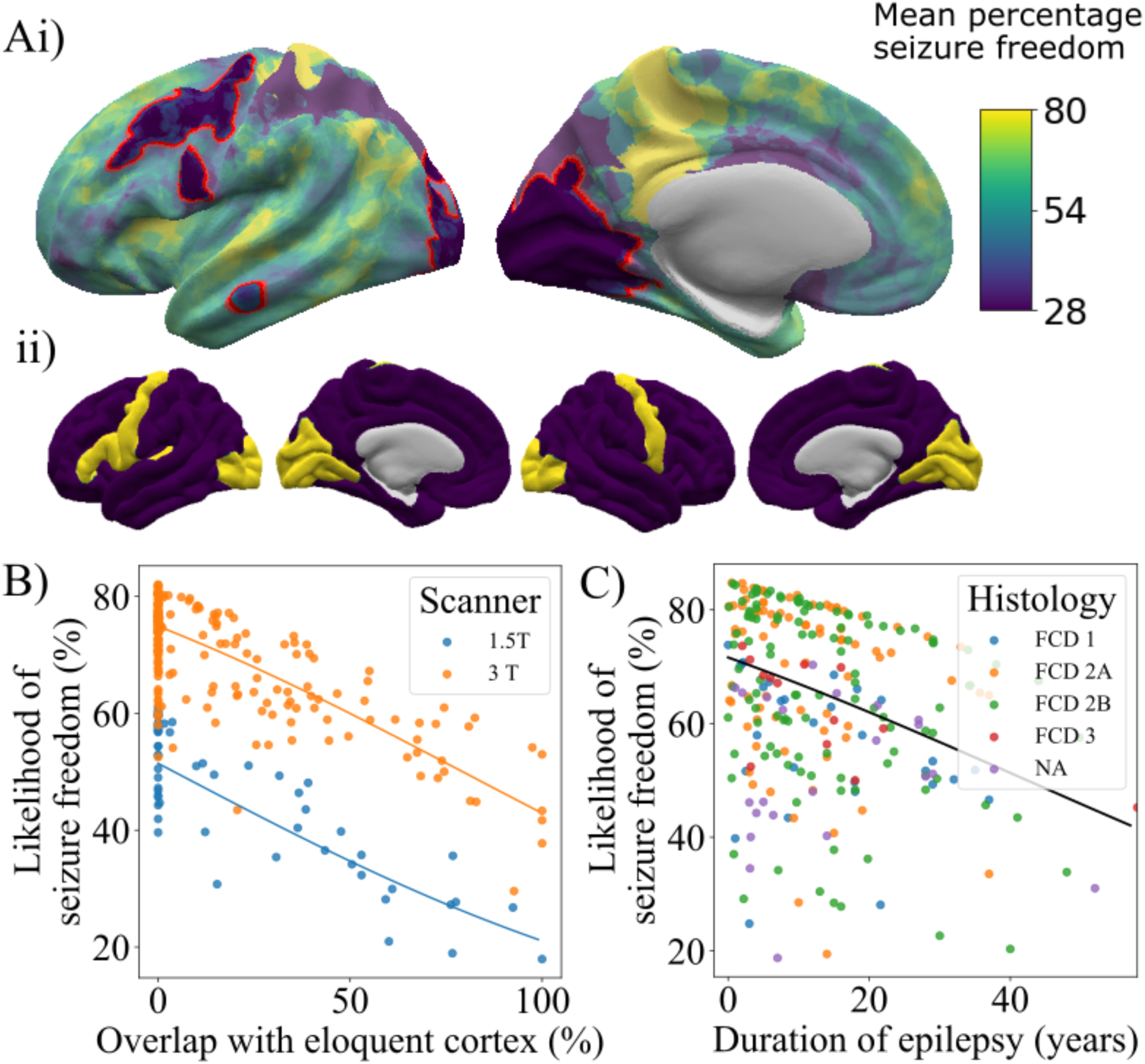
Effect of lesion location, duration of epilepsy and histopathological subtype on seizure freedom. (**Ai**) Percentage of patients seizure free (%) according to lesion location across the cerebral cortex. Visual, motor and premotor areas had a low percentage of seizure free patients (30-40%). (**Aii**) Mask of eloquent cortex. (**B**) Impact of overlap of lesion with eloquent cortex and MRI scanner field strength on likelihood of seizure freedom. (**C**) Impact of duration and histopathology on predicted percentage likelihood of seizure freedom.

The second logistic regression model was fitted with duration of epilepsy, age of epilepsy onset, MRI-negative status, scanner, lesion overlap with eloquent cortex and lesion size. Seizure freedom decreased as the overlap with eloquent cortex increased (Fig. 3B, *β* = −1.39, *p* < 0.001), with a 50% overlap between the lesion and eloquent cortex associated with a 16% decrease in the likelihood of seizure freedom compared to no overlap. Being scanned on a 1.5T MRI scanner was associated with lower likelihood of seizure freedom, approximately 25% lower (Fig. 3B, *β* = 1.04, *p* < 0.001). Likelihood of seizure freedom decreased with longer duration of epilepsy (Fig. 3C, *β* = −1.23, *p* < 0.01), with a 10-year increase in duration of epilepsy associated with a 5% decrease in likelihood of seizure freedom. There was no significant association between age of epilepsy onset, lesion size or MRI-negative status and postsurgical seizure freedom. However, the directions of the effects were in keeping with previous literature, where larger lesions (*β* = −0.66) and having been reported MRI-negative (*β* = −0.32) tended towards worse outcomes. In the separate model, including age at MRI scan, seizure freedom decreased as age at MRI scan increased (*β* = −1.07, *p* < 0.01).

The third logistic regression model additionally included histopathological diagnosis and an interaction term between histopathological diagnosis and lesion size. There was no significant association between FCD subtype and seizure freedom, nor was there an interaction between FCD subtype, lesion size and seizure freedom. Duration of epilepsy, scanner field strength and overlap with eloquent cortex remained significant.

10-fold cross-validation of the second logistic regression model including only presurgical variables was used to calculate the predictive value of the model. The model for predicting seizure freedom had a sensitivity of 92%, specificity of 30%, positive predictive value of 70% and negative predictive value of 67%.

### 5. Inter-relationships between demographic, lesional and surgical variables

Fig. 4A displays significant relationships between demographic, lesional and surgical variables after systematic evaluation of all inter-relationships. Full statistics are reported in Supplementary Fig. 3. Results of interest have been highlighted below.

**Figure 4.**
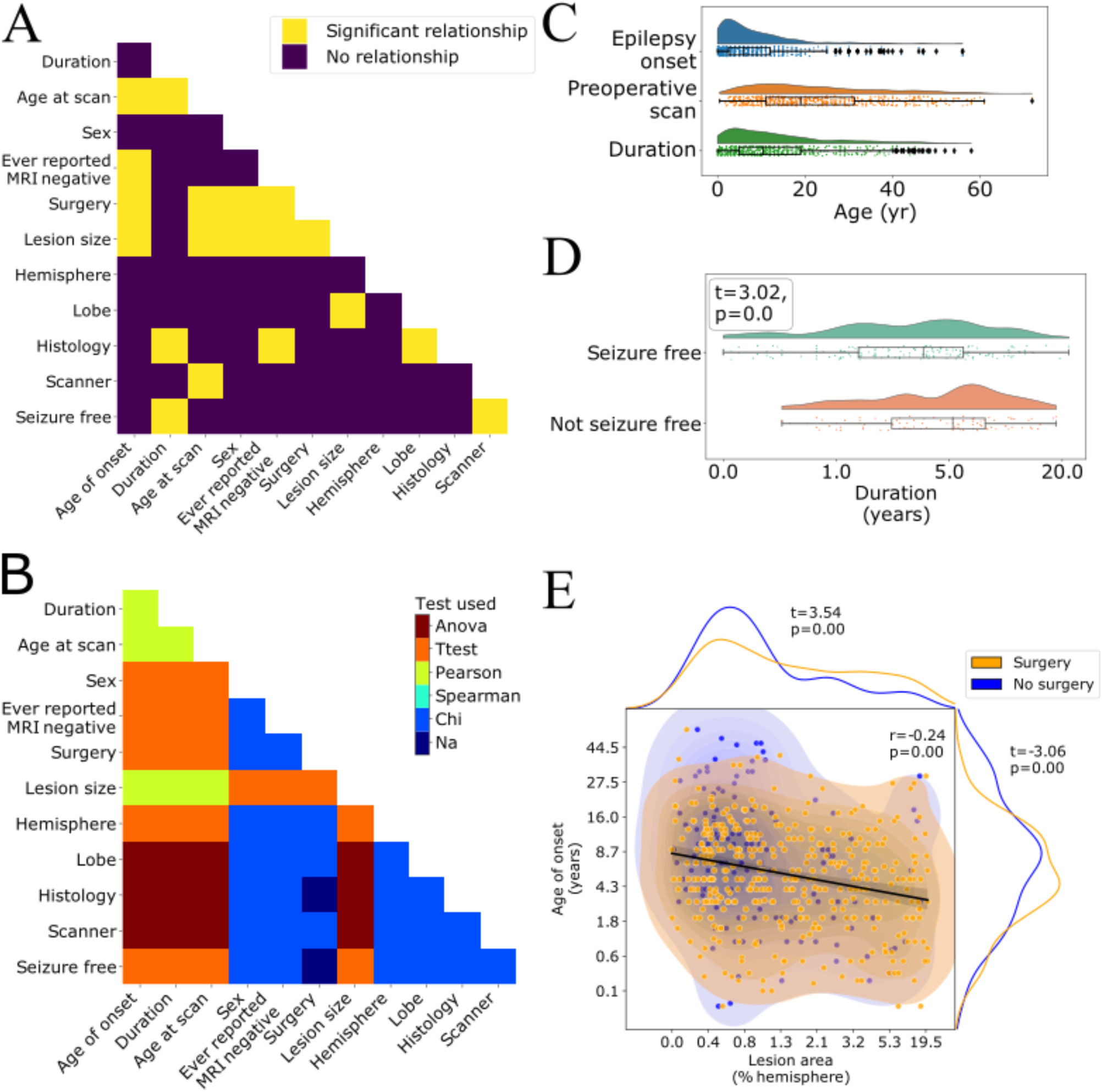
Interrelationships between features. (**A**) Pairwise comparison of demographic and clinical features. Significant relationships after correction for multiple comparisons are shown in yellow. (**B**) Statistical test used for each pairwise comparison. (**C**) Distributions of age of epilepsy onset, age at MRI scan and duration of epilepsy. (**D**) Duration of epilepsy is significantly associated with seizure freedom (*t* = −3.0, *p* < 0.001). Patients with longer durations of epilepsy are less likely to be seizure free. (**E**) Age of epilepsy onset, lesion size (as a percentage of the total hemisphere size) and seizure freedom are significantly associated. Larger lesions are associated with younger age of epilepsy onset (*r* = −0.24, *p* < 0.001) and are more likely to be operated (*t* = 3.69, *p* < 0.001). Similarly, patients with a younger age of epilepsy onset are more likely to be operated (*t* = −3.76, *p* < 0.001).

#### Relationships with age at MRI scan

The distributions of age at epilepsy onset, age at MRI scan and duration of epilepsy revealed that overall there was a long interval between patients developing epilepsy and having their MRI scan (Table 1, Fig. 4C). While 68% of patients have epilepsy onset before age 10, 51% of patients wait over 10 years before having their MRI scan and consequently undergoing presurgical evaluation. There was a small but significant negative correlation between age at scan and lesion surface area (*r* = −0.19, *p* < 0.001), i.e., patients with larger lesions had presurgical evaluation younger. Patients scanned with 1.5T MRI were on average younger than those with 3T (*F* = 8.58, *p* < 0.001). Lastly, older patients were less likely to have had surgery (*t* = −3.76, *p* < 0.001).

#### Relationships between lesion size, histopathology, surgery and ever reported MRI-negative

Patients with earlier epilepsy onset had larger lesions (*r* = −0.24, *p* < 0.001, Fig. 4E), were less likely to have ever been reported MRI-negative (*t* = −3.70, *p* < 0.001) and more likely to have had epilepsy surgery (*t* = −3.38, *p* < 0.01, Fig. 4E). Patients with larger lesions were more likely to have had surgery (*t* = 3.69, *p* < 0.001, Fig. 4E). FCD Type IIA were more likely to have been MRI-negative than FCD Type IIB (*χ*^*2*^ = 12.2, *p* < 0.01, Tukey’s post-hoc, *p* < 0.01).

#### Relationships with lesional lobe

Lesion surface area was significantly associated with lobe (*F* = 6.9, *p* < 0.001), driven by temporal lobe lesions being larger than frontal (Tukey’s post-hoc *p* < 0.01) and parietal lesions (Tukey’s post-hoc *p* < 0.05). FCD Type I and III lesions were more frequently located in the temporal lobe (*χ*^*2*^ = 70.2, *p* < 0.001, Tukey’s post-hoc *p* < 0.001, Fig. 1E).

#### Relationships with seizure freedom

Patients with longer durations of epilepsy were less likely to be seizure free (*t* = −3.0, *p* < 0.001). Patients with 1.5T MRI scans were less likely to be seizure free than those with 3T imaging (*χ*^*2*^ = 15.7, *p* < 0.001). No other factors survived correction for multiple comparisons (Supplementary Fig. 3).

## Discussion

In this multi-centre study of 580 patients with FCD, lesions were non-uniformly distributed across the cerebral cortex, with predominance in the superior frontal sulcus, frontal pole and temporal pole. Vertex-specific lesion location was significantly associated with age of epilepsy onset, duration of epilepsy, age at MRI scan and lesion size. Likelihood of seizure freedom post-operatively varied considerably according to lesion location, with lesions in visual, motor and premotor areas associated with much lower rates of seizure freedom than elsewhere, likely attributable to neurosurgical caution in resecting lesions around eloquent cortex. A model incorporating overlap with eloquent cortex alongside duration of epilepsy, age at epilepsy onset, MRI scanner field strength, whether the patient was ever reported MRI negative and lesion size had a positive predictive value of 70% and negative predictive value of 67%.

The atlas of FCD lesion location highlighted a non-uniform distribution across and between cortical lobes (Fig. 1). It substantiated previous studies finding that FCDs were more common in frontal (particularly FCD Type II) and temporal (FCD Types I & III) lobes.^3,4^ Additionally, this atlas extends previous understanding, demonstrating “hot-spots” in the superior frontal sulcus driven by FCD Type IIB, and a cluster of temporal pole lesions in all FCD subtypes. Understanding of sites of predilection for FCD are useful in the clinical search for FCDs as well as for radiology teaching. For research purposes, the atlases highlight areas where improvements in imaging could lead to improved FCD detection. For instance, high-field MRI has difficulties with signal inhomogeneities in the temporal lobes, an area of high-incidence for these subtle lesions and therefore a critical area for improvement. Furthermore, regional variations in FCD incidence provide directions for future research including whether the somatic mosaic mutations causing these malformations of cortical development occur non-uniformly or whether yet unidentified neurobiological properties determine their epileptogenicity.

Linking individual clinical and demographic data with lesion location uncovered relationships between age of epilepsy onset, duration of epilepsy, lesion size and lesion location. Lesions in primary sensory, motor and visual areas were associated with earlier epilepsy onset, while lesions in higher-order association cortex were associated with later epilepsy onset (Fig. 2A & E). This relationship may reflect differing developmental trajectories of these areas,^22,23^ with seizures initiating as a result of the development of particular cortical properties. However, differing seizure semiologies may also be a contributing factor, where more subtle seizure symptoms are not attributed to a diagnosis of epilepsy for longer.

Most patients had epilepsy onset during childhood (median onset = 6.0 years), but the median age at MRI scan was 19.0 years (Table 1), indicating many patients had long delays between epilepsy onset and potentially curative epilepsy surgery (Fig. 4C, median duration = 10.4 years). Longer duration of epilepsy is associated with increased morbidity, mortality and with worse postsurgical outcome (Fig. 3B & 4A).^4,24^ In our cohort, patients with a longer duration of epilepsy were more likely to have lesions in the frontal cortex, particularly around the central sulcus. Factors contributing to longer duration of epilepsy might include diagnostic and surgical challenges such as lesion conspicuity, MRI scanning protocol and whether a lesion is in eloquent cortex. In keeping with this, larger lesions were more likely to have been operated (Fig. 4D). Other reasons for prolonged durations of epilepsy might include trials of anti-epileptic drugs, alongside under and delayed referral to specialist epilepsy centres.^25^

Consistent with a recent study,^4^ 66% of patients with FCD in the MELD cohort were seizure free postsurgically. We found that a longer duration of epilepsy was significantly associated with a reduced chance of seizure freedom (Fig. 4A), but the impact of duration was small, with a 5% decrease in likelihood of seizure freedom for every 10 year increase in duration of epilepsy (Fig. 3C). By contrast, there was a striking effect of lesion location on seizure freedom, with the likelihood of seizure freedom dropping from 70% when lesions had no overlap with eloquent areas to 54% when there was 50% overlap, representing a 16% decrease (Fig. 3B). The most likely reason is that neurosurgical resection plans were intentionally cautious in an attempt to avoid resecting motor and visual cortex or the adjacent white matter tracts such as the optic radiation or corticospinal tracts to minimise the risk of deficits such as hemiparesis or hemianopia.^26^ Additionally, the medial occipital cortex is located on the medial wall of the cortex, restricting ease of surgical access. Both factors could contribute to higher rates of incomplete resections of the dysplastic tissue known to be associated with worse postsurgical outcome.^11^

Statistically significant factors are not necessarily useful predictors.^27^ However, in this case, the predictive model of postoperative seizure freedom, including lesional overlap with eloquent cortex alongside duration of epilepsy, age of epilepsy onset, MRI negative status, MRI scanner field strength and lesion size, demonstrated predictive power on unseen data. As such these tools can help inform presurgical decision making and more broadly can be integrated with other established clinical predictors to improve predictive modelling of seizure freedom.^28,29^

One strength of our study is the inclusion of both operated and non-operated patients. This helped to minimise ascertainment bias in the dataset, as the lesional distribution of non-operated patients captured more lesions in eloquent language areas (e.g. left inferior frontal gyrus) or cortex which is more surgically challenging to resect (e.g. insula, Supplementary Fig. 2), while purely postsurgical cohorts may undersample these patients. However, it is important to note that our cohort primarily consisted of patients with epileptogenic, drug-resistant FCDs. Independent studies are needed to establish the distributions of non-epileptogenic or drug-responsive FCDs which may not present to epilepsy surgery centres.

One limitation is the number of clinical and MRI variables collected. Future work including more detailed clinical information, such as seizure types, seizure burden, electrophysiology, medication, genetics and extent of lesion resection from postoperative MRI scans might further our understanding of FCD and enable improved predictive models for seizure freedom. Additionally, the clinical variable “ever reported MRI-negative” is imperfect, depending on MRI protocols and the expertise of radiological review. Nevertheless over a large sample size, it does indicate that some lesions were more subtle. From our data FCD Type IIB was less likely to be MRI-negative, and therefore there may be an ascertainment bias in our dataset, with an overrepresentation of FCD Type IIB.

As a subtle pathology on MRI, manual masking of FCD lesions is challenging. There is likely to be some heterogeneity in the masking process. At each site lesions were delineated by different individuals. Additionally, for some lesions that were difficult to detect on MRI delineation of the lesion boundaries was guided by postsurgical resection cavities. By contrast, other lesions might extend beyond what was visible. Future studies using automated lesion detection and masking^7^ may yield more observer-independent and consistent lesion masks. Nevertheless, the stability of the lesion distribution pattern when randomly subsampling the cohort (Fig. 1C) indicates that the core findings of this study are robust to idiosyncrasies of particular lesion masks.

Large collaborative initiatives have shown the power of big data to answer clinically relevant questions.^9,30,31^ Here open-science practices enabled mapping of the topographic distribution of epileptogenic FCDs across the cerebral cortex, a departure from the coarse, lobar annotations usually described. The surface and volumetric atlas of lesion location will serve as a diagnostic and teaching aid in the radiological search for an individual patient’s subtle lesion. The predictive models of postsurgical seizure freedom can be used to estimate individual postsurgical seizure freedom based on a lesion’s proximity to eloquent cortex. This could also inform presurgical decision making, resection planning and risk counselling of patients. Lastly, through making all our code available and providing user-friendly interactive notebooks detailing how to run all our analyses, this analysis framework can be replicated by researchers in epilepsy and other focal neurological conditions.

## Data Availability

Code to reproduce all analyses and figures in this manuscript is available to download from github.com/MELDproject. Requests can be made for access to the full MELD dataset.

## Abbreviations

FCD: Focal cortical dysplasia
MELD: Multicentre Epilepsy Lesion Detection

alphabetical order, separated by semi-colons; 12 point; abbreviations should be in the format: UPDRS = Unified Parkinson’s disease Rating Scale

The following do not need defining: AIDS; ANOVA; ATP; cDNA; CNS; CSF; CT; DNA; ECG; EEG; ELISA; EMG; GABA; HIV; MRI; PBS; PCR; PET; REM; RNA; mRNA; tRNA

## Supplementary material

‘Supplementary material is available at *Brain* online’.

## Figure legends

**Supplementary Figure 1.**
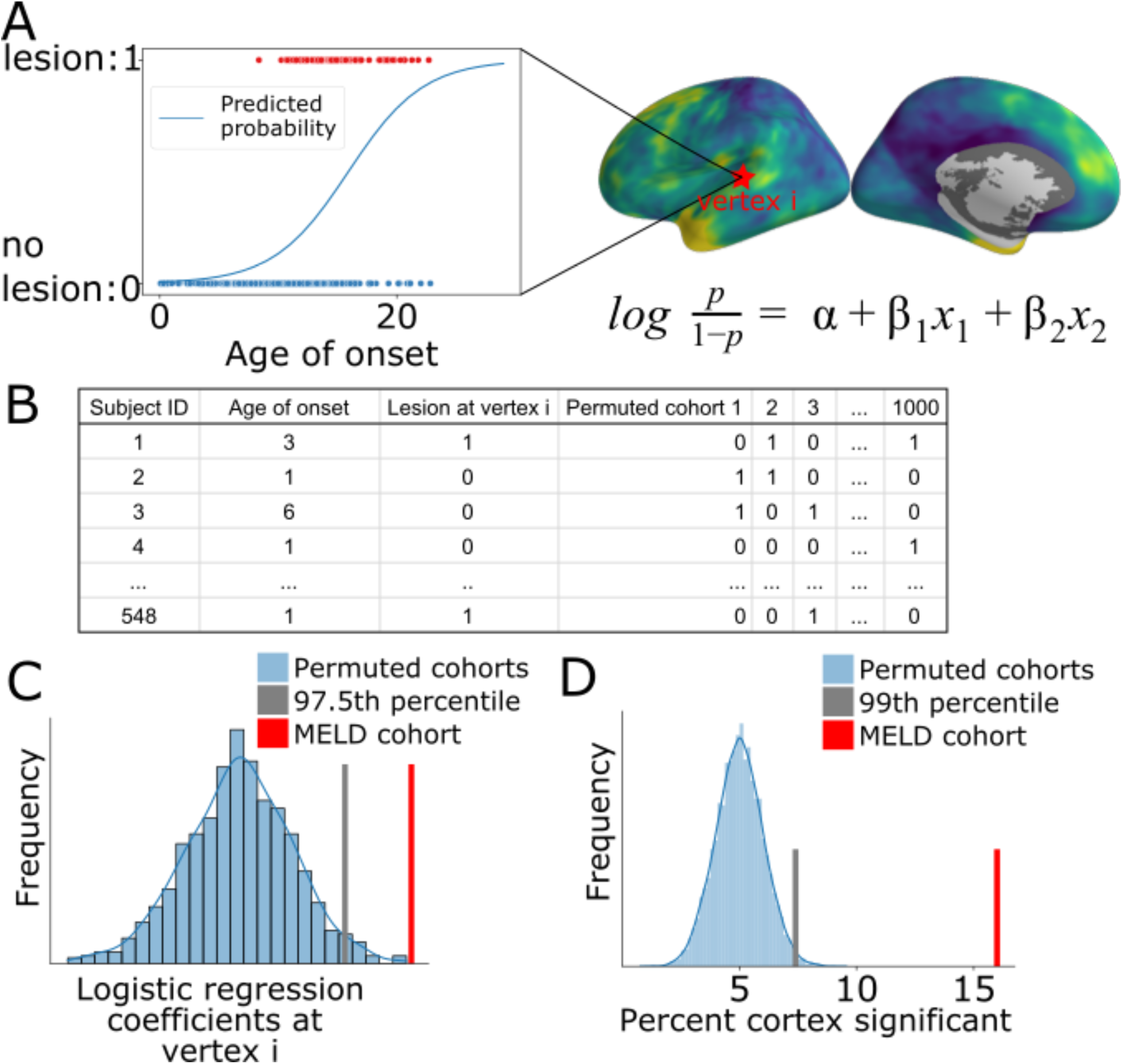
Overview of logistic regression framework to identify presurgical predictors of lesion location. (**A**) Logistic regression model including age of epilepsy onset, sex, ever reported MRI-negative, duration of epilepsy and lesion hemisphere was fitted to predict the presence / absence of a lesion at every vertex. (**B**) 1000 permuted cohorts created where presence / absence of a lesion at a particular vertex is random. (**C**) Normal distribution of coefficients from permuted cohorts at vertex i. The coefficient from actual data (red) is greater than 97.5% of the coefficients from the permuted cohorts, and therefore vertex i is considered significant. (**D**) Normal distribution of the percentage of vertices that were significant for a particular factor (e.g., age of onset) in the permuted cohorts. The factor (red) was considered significantly related to lesion location as the number of significant vertices exceeded the 99th percentile value from the permuted cohorts.

**Supplementary Figure 2.**
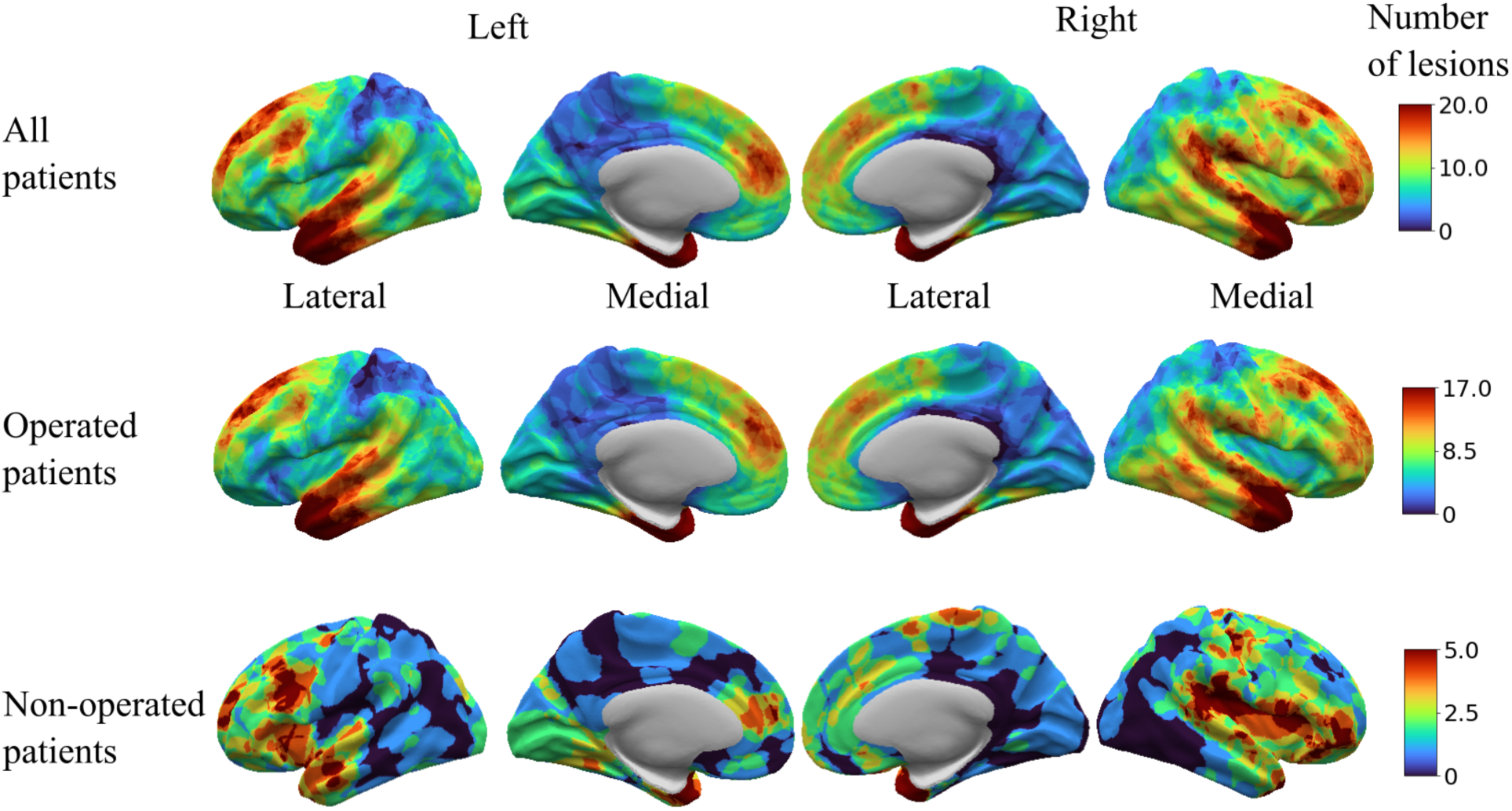
Bilateral distributions of FCDs. Distributions of FCD lesions on left and right hemispheres in the whole MELD cohort, patients who had undergone surgery and those that had not. The pattern in the whole and operated cohorts appear symmetric, whereas patients who had not undergone surgery appeared to have more lesions in the left inferior frontal gyrus, near Broca’s area. Additionally lesions in the non-operated cohort appear to be more frequently located in the insula, a diagnostically and surgically challenging area for cortical resection.

**Supplementary Figure 3.**
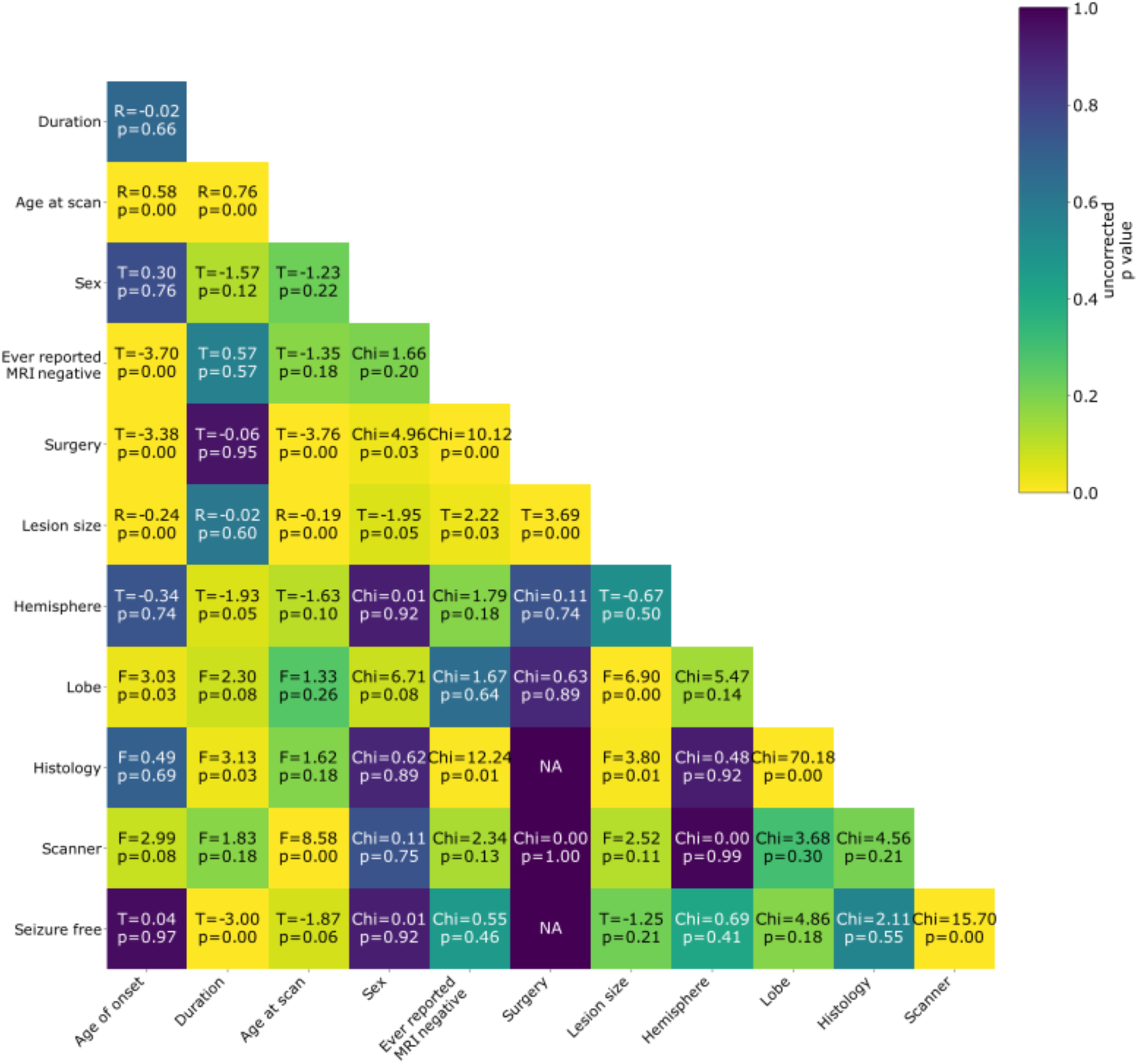
Pairwise comparisons between all demographic and clinical features. Test statistics and uncorrected p values are displayed. See Figure 4 for a more detailed exploration of findings.

